# Towards a simulation framework for optimizing infectious disease surveillance: An information theoretic approach for surveillance system design

**DOI:** 10.1101/2020.04.06.20048231

**Authors:** Qu Cheng, Philip A. Collender, Alexandra K. Heaney, Xintong Li, Rohini Dasan, Charles Li, Joseph A. Lewnard, Jon Zelner, Song Liang, Howard H. Chang, Lance A. Waller, Benjamin A. Lopman, Changhong Yang, Justin V. Remais

## Abstract

Infectious disease surveillance systems provide vital data for guiding disease prevention and control policies, yet the formalization of methods to optimize surveillance networks has largely been overlooked. Decisions surrounding surveillance design parameters—such as the number and placement of surveillance sites, target populations, and case definitions—are often determined by expert opinion or deference to operational considerations, without formal analysis of the influence of design parameters on surveillance objectives. Here we propose a simulation framework to guide evidence-based surveillance network design to better achieve specific surveillance goals with limited resources. We define evidence-based surveillance design as a constrained, multi-dimensional, multi-objective, dynamic optimization problem, acknowledging the many operational constraints under which surveillance systems operate, the many dimensions of surveillance system design, the multiple and competing goals of surveillance, and the complex and dynamic nature of disease systems. We describe an analytical framework for the identification of optimal designs through mathematical representations of disease and surveillance processes, definition of objective functions, and the approach to numerical optimization. We then apply the framework to the problem of selecting candidate sites to expand an existing surveillance network under alternative objectives of: (1) improving spatial prediction of disease prevalence at unmonitored sites; or (2) estimating the observed effect of a risk factor on disease. Results of this demonstration illustrate how optimal designs are sensitive to both surveillance goals and the underlying spatial pattern of the target disease. The findings affirm the value of designing surveillance systems through quantitative and adaptive analysis of network characteristics and performance. The framework can be applied to the design of surveillance systems tailored to setting-specific disease transmission dynamics and surveillance needs, and can yield improved understanding of tradeoffs between network architectures.

**Author summary:** Disease surveillance systems are essential for understanding the epidemiology of infectious diseases and improving population health. A well-designed surveillance system can achieve a high level of fidelity in estimates of interest (e.g., disease trends, risk factors) within its operational constraints. Currently, design parameters that define surveillance systems (e.g., number and placement of the surveillance sites, target populations, case definitions) are selected largely by expert opinion and practical considerations. Such an informal approach is less tenable when multiple aspects of surveillance design—or multiple surveillance objectives— need to be considered simultaneously, and are subject to resource or logistical constraints. Here we propose a framework to optimize surveillance system design given a set of defined surveillance objectives and a dynamical model of the disease system under study. The framework provides a platform to conduct *in silico* surveillance system design, and allows the formulation of surveillance guidelines based on quantitative evidence, tailored to local realities and priorities. The approach facilitates greater collaboration between health planners and computational and data scientists to advance surveillance science and strengthen the architecture of surveillance networks.

## 1 Introduction

Infectious disease surveillance systems provide vital information on patterns of disease occurrence across space, time, and populations of interest, and ultimately provide the basis for evidence-based disease control policy decisions [1]. Considerable progress has been made supporting infectious disease control decision-making with computational approaches to evaluate the outcomes of alternative decisions [2]. Examples include optimizing when, where, and among which populations to allocate public health resources [3, 4], determining the optimal balance between multiple intervention approaches (e.g., case detection, treatment, vaccination, and sanitation improvement) [5-8], and optimizing the start time, duration, and dose of drug treatment programs [9, 10]. In contrast, little attention has been paid to the development of tools for improving infectious disease surveillance system designs, and formalization of methods to optimize surveillance networks has largely been overlooked.

The ‘design parameters’, which are the high-level characteristics that define infectious disease surveillance networks—such as the number and locations of surveillance sites, sampling frequency for laboratory testing or community-based surveys, and selection of diagnostic techniques—can greatly influence the degree to which the resulting surveillance data serves public health objectives, including early detection of outbreaks (e.g., the coronavirus disease outbreak in 2020) [11], improved understanding of disease emergence and spread [12], and accurate measurement of the impact of interventions [13]. Thus, key design parameters can be modified in a manner informed by optimization analysis such that the system better achieves specific surveillance goals. Examples include relocating and adding reporting sites to predict the temporal trend of diseases more accurately [14]; changing diagnostic approaches/case definitions to increase the chance of detecting cases [15]; and targeting of sampling towards specific subpopulations to improve the timeliness of outbreak detection [16, 17].

In practice, surveillance system design parameters are often set in an *ad hoc* fashion based on operational considerations (e.g., budget, convenience, political agendas), rather than through quantitative evaluation of how alternative designs might impact surveillance system objectives. For instance, World Health Organization (WHO) recommends selection of influenza surveillance sites based on the facilities’ willingness to participate, availability of necessary laboratory and information infrastructure, ability to cover the surveillance cost, and representativeness of the general population. Notably absent from these criteria is the degree to which the network’s performance on specific surveillance objectives will be enhanced [13]. The absence of objective criteria and methods to evaluate and iteratively reconfigure surveillance system design can lead to inefficient use of limited resources. For example, in China, current requirements specify that 5-15 influenza-like illness (ILI) cases are required to be sampled per week at each of the 556 influenza sentinel hospitals for laboratory confirmation [18]. If the total sample size is fixed, it may be that reducing the number of sentinel sites (e.g., prioritizing sites in populous regions and with high levels of population movement), while increasing the sample sizes at the remaining sites, could yield more timely detection of outbreaks with the same level of resources. What is more, because disease surveillance systems generally operate in pursuit of multiple objectives, decision-making surrounding optimal design can be highly counterintuitive.

Recent research has provided some early examples of quantitative infectious disease surveillance design optimization [19, 20]. In one study, researchers estimated that an optimal relocation of Iowa’s existing 22 ILINet sentinel sites could increase population coverage of the network from 56% to 75% [21]. As another example, targeted surveillance of pregnant women over blood donors for compulsory diagnostic testing was estimated to increase the weekly probability of detecting at least one Zika case from 11% to 40% [15]. While these and other studies serve as foundational examples, the methods utilized in these analyses are targeted towards narrow, study-specific objectives and specific networks, and are challenging to generalize to other—even closely related—surveillance design optimization problems. What is more, prior studies have not attempted to articulate a general theory of surveillance design optimization and decision-making.

Here, we present for the first time a unified analytical framework for quantitative infectious disease surveillance system optimization, accommodating multiple surveillance design parameters, objectives, operational constraints, and underlying disease processes. A common framework and standard terminology can enable closer collaboration between and among computational researchers, public health officials, and other stakeholders regarding the design and implementation of infectious disease surveillance systems. This in turn can accelerate the pace of methodological innovations and facilitate the development of surveillance design theories that anticipate and respond to current and future epidemiological challenges. Furthermore, a generalized framework can inspire the application of quantitative surveillance optimization across broader settings, resulting in system designs better aligned with local realities and public health priorities.

## 2 Surveillance design as a multi-objective, multi-dimensional, constrained and dynamic optimization problem

The search for optimal disease surveillance designs is a highly complex problem. This results from the multiple, often competing goals of surveillance data collection, idiosyncratic surveillance network design, the need to represent operational constraints that govern surveillance systems, and the complexity and dynamic nature of diseases under surveillance. Simple optimization problems involving a single design parameter and objective for a given target disease—such as the optimal placement of a new surveillance site to maximize the proportion of influenza cases detected—may be solved in relatively straightforward fashion by testing all possible designs and choosing the design that generates optimal network performance (e.g., the new site location that results in the highest proportion of cases detected overall). However, surveillance network optimization quickly becomes non-trivial when the design space increases (e.g., selecting 10 sites out of 200 alternative sites), when multiple objectives (such as increasing case detection, improving spatial and temporal trend coverage, and risk factor identification) are subject to simultaneous analysis and optimization, or when optimization is subject to constraints regarding resource limitations and operational plausibility. Uncertainty regarding the functioning of the epidemiologic system and shifts in patterns of diseases further complicate matters. Hence, our optimization goals are multidimensional, dynamic, and stochastic. In this section, we describe the relevance of surveillance objectives, network design parameters, operational constraints and dynamic disease systems to the pursuit of surveillance optimization.

### Multiple objectives

Disease surveillance systems are established and designed for diverse purposes, including to collect data to understand variations in disease frequency across populations, space, and time, to monitor pathogen composition over time, to detect outbreaks and forecast epidemics, to assess the impact of interventions, and to determine risk factors associated with diseases. Most surveillance systems operate with multiple public health objectives and multiple logistical constraints. Hence, surveillance system designs should generally be subject to multi-objective optimization, and tradeoffs between different objectives must be considered. For instance, if the goals of a system are to both estimate prevalence and assess the impact of risk factors, the network design should be subjected to optimization that considers both objectives using a framework that is capable of capturing tradeoffs between designs with respect to achieving the two objectives.

### Multiple design parameters

Surveillance system structure and design can be decomposed into a multitude of characteristics, operational details, and features that influence the performance of surveillance networks. Design parameters can lend themselves to representation and simulation within models. Multiple design parameters may be amenable to optimization analysis, either through single or multivariate optimization. For example, to improve estimation of disease incidence, either the accuracy of diagnostics at existing reporting facilities or the number of facilities in the reporting network, or both, can be modified. Other design parameters, such as when, where, and among which populations to implement targeted sampling efforts may also be entered into the analysis, greatly expanding the dimensionality of the problem. Moreover, the set of design parameters to optimize depends on the surveillance goals. For example, when the surveillance goal is accurate estimation of the temporal trend of a disease, it may be that the placement of sites is less important than sampling frequency. Table 1 shows examples of design parameters from select real world infectious diseases surveillance systems, and their potential impacts on surveillance system performance.

**Table 1.** Example surveillance system design parameters and their potential impacts on surveillance performance.

| Design parameter | Definition | Potential impacts on surveillance performance | Example designs | Example surveillance system | Ref. |
| --- | --- | --- | --- | --- | --- |
| **Target population** | Population to be monitored for outcomes of interest. | Target populations representative of a general population provide a means of tracking overall disease incidence and trends in the population as a whole. Target populations informed by demographic differences in disease risk, transmission potential, or detection probability may provide advantages for monitoring outcomes in vulnerable populations, anticipating outbreaks, or tracking rare diseases. | All persons >2 years of age residing in homes | Republic of South Africa HIV prevalence survey | [22] |
|  |  |  | Pregnant women and infants | US Zika Pregnancy and Infant Registry | [23] |
| **Site enrollment** | The inclusion of hospitals and other facilities in passive reporting networks, or selection of locations for active or environmental surveillance | Site selection influences factors such as population coverage and representativeness, diagnostic quality, the speed at which spreading outbreaks may be detected, and informational redundancy due to spatial proximity or other sources of similarity between locations | Hospitals in Maluku, North Sulawesi, East Kalimantan, North Sumatra, Yogyakarta and West Nusa Tenggara | Indonesia influenza sentinel surveillance system | [24] |
|  |  |  | Health centers in Dembi, Asendabo, Tulubolo, Guangua, Bulbula, Dhera, Welenchity, Metahara, Asebot, and Kersa | Ethiopia malaria sentinel surveillance | [25] |
| **Sampling strategy** | Type of sampling used to identify cases among the target population. | Sampling strategies influence the representativeness of surveillance data, as well as the ability of surveillance systems to detect rare or underreported conditions. Strategies that adequately characterize a general population may be biased with respect to critical subpopulations, especially those facing stigma. | Hospital-based convenience sampling (e.g. every fourth patient meeting case definition) | Bangladesh rotavirus surveillance system | [26] |
|  |  |  | Respondent-driven sampling, which uses existing samples in high-risk groups (e.g., intravenous drug user, men who have sex with men) to recruit new samples, then uses a model to correct for potential bias in the nonprobability sampling | Central America sexual behaviors and HIV prevalence survey | [27] |
| | | | Multistage cluster sampling, which selects districts first according to their population size, then selects communes within the selected districts by simple random sampling, and selects all residents aged $\geq 15$ years in the selected communes | Viet Nam national survey of tuberculosis prevalence | [28] |
| **Sampling intensity** | Number of samples per sampling interval | Under operational constraints, the choice between sampling more frequently but with low intensity or less frequently with higher intensity represents a tradeoff between the ability to resolve high frequency changes in outcomes of interest, or timeliness of detection, and reducing statistical uncertainty | 3 adults and 2 children per week | Malaysia laboratory-based influenza surveillance system | [29] |
|  |  |  | 5 mild cases serotyped per month per site | China hand foot mouth disease sentinel surveillance system | [30] |
| **Sampling seasonality** | Pre-determined changes in sampling intensity over time | Year-round sampling increases the chances of detecting unexpected changes in disease incidence. However, if disease seasonality is static and well-understood, resources may be better used for intensive seasonal sampling | Year-round | New Zealand virological surveillance system | [31] |
|  |  |  | Transmission season (June-October) | China dengue virological surveillance system | [32] |
| **Laboratory diagnostics** | Methods used to determine the presence of a pathogen or syndrome of interest. | Diagnostic tests and other related factors such as specimen types, the quality of the specimen, and the time from onset to specimen collection can influence the sensitivity and specificity of the surveillance system. | Isolation of *Bordetella pertussis* from clinical specimen and/or a four-fold or greater increase in titer of antibody against *B. pertussis* between acute and convalescent sera | China pertussis surveillance system | [33] |
|  |  |  | Isolation of *B. pertussis* from clinical specimen and positive polymerase chain reaction (PCR) for *B. pertussis* | US CDC pertussis surveillance system | [34] |
| **Case definition** | Diagnostic criteria to classify outcomes of interest. | Case definitions can influence factors such as the severity and characteristics of cases identified, and the sensitivity and specificity of the system. | Influenza-like illness, defined as an acute respiratory infection with measured fever of $\geq 38^{\circ}\text{C}$ and cough with onset within the last 10 days | WHO global influenza surveillance | [35] |
| | | | Severe acute respiratory infection, defined as an acute respiratory infection with history of fever or measured fever of $\geq 38^{\circ}\text{C}$ and cough with onset within the last 10 days and requires hospitalization | | |

### Operational constraints

Operational restrictions on surveillance system designs—due to budgetary, logistical, political and cultural considerations—add critical constraints to the optimization problem. Absent constraints, the optimal design may be self-evident, e.g., sampling at maximal frequency and intensity. Yet when there is a fixed budget for samples, the optimal balance between design parameters—say, number of samples and sampling frequency— depends on the relative value of precise cross-sectional estimates of disease prevalence versus characterizing disease incidence over time, which in turn depends on the specific objectives of surveillance and the dynamics of the underlying disease system.

### Dynamic and imperfectly understood disease systems

Surveillance systems must respond to shifts in the epidemiology of target infections. Optimal designs will likely shift in response to the evolution of underlying epidemiology and available knowledge. For instance, as infections emerge, become endemic, or approach elimination within populations or subpopulations, the goals of surveillance, and the resulting optimal designs, can (and must) evolve alongside them. The dynamic nature of optimal surveillance design may be especially important in emerging economies that are undergoing epidemiologic transitions. For instance, as a region or nation approaches elimination of a particular infectious disease, surveillance goals generally shift from enumeration of endemic cases occurring in the general population to detection of nexuses of sporadic transmission. This may require new designs (e.g., shifting to a more sensitive diagnostic test within a limited area, or increasing the coverage of subpopulations involved in ongoing transmission), and adjustment of system objectives (e.g., maximize detection of the few remaining cases instead of minimizing false positive detections). Additionally, as cases caused by novel pandemics (e.g., the 2020 coronavirus disease pandemic, or 2009 H1N1 pandemic) start to increase exponentially, surveillance systems may need to switch from tracking individual cases to population-based surveillance (e.g., pathogen testing for a proportion of patients with a non-specific syndrome) in order to monitor the progression of the outbreak and develop mitigation strategies without depleting public health resources.

## 3 A framework for surveillance simulation and optimization

The aforementioned challenges of surveillance optimization—multiple objectives, combinatorial complexity of relevant design parameters, operational constraints, and dynamic and uncertain epidemiology of target diseases—suggest the need for a generalized framework for surveillance network optimization. Advances in computation for simulation-based studies have benefitted many related fields, including optimal disease control [36-39], yet applications of simulation optimization to the design of disease surveillance networks have scarcely been pursued. In the following sections, we detail a simulation and optimization framework for designing infectious disease surveillance networks, and demonstrate its application in a site selection context. Our framework facilitates a quantitative approach to designing surveillance systems tailored to local disease transmission dynamics and surveillance needs, as well as a more general study of optimal network design principles under varying objectives and epidemiological circumstances.

Broadly, our framework (Figure 1) allows for evaluation of surveillance system performance across a predefined design space under different epidemiologic scenarios (disease system model) and network characteristics (surveillance model). Numerical optimization algorithms are applied to efficiently identify the region(s) of design space that yield superior network performance based on one or more specific surveillance goals (simulation optimization search). The optimization procedure (Figure 1 and Box 1) yields a set of network designs (i.e., optimal design parameter values) that maximize performance with respect to the specified public health goal(s), according to the specified data and models.

**Figure 1.**
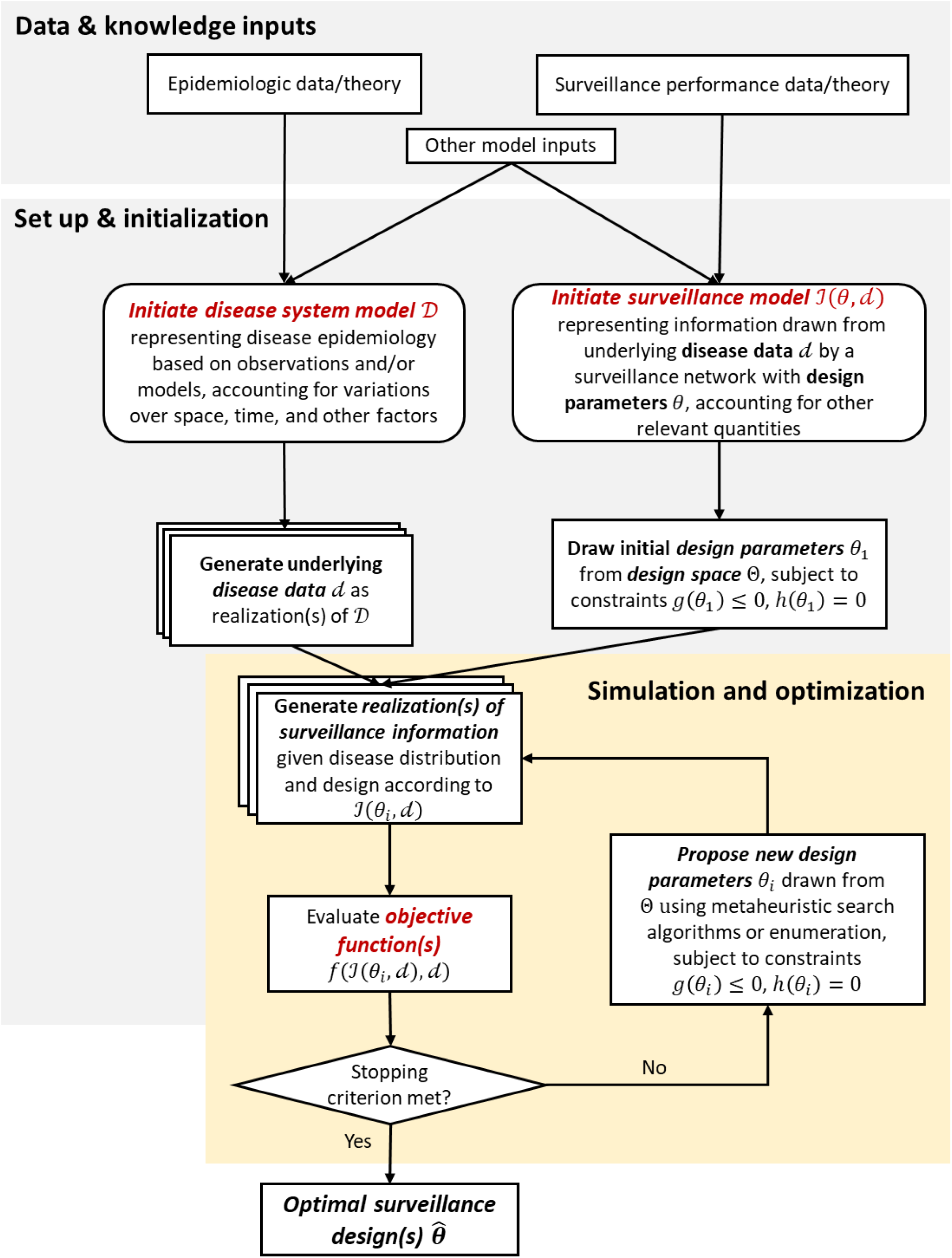
Schematic of surveillance system optimization. The surveillance system optimization procedure uses data and knowledge about disease transmission and case ascertainment to identify optimal surveillance designs with regard to predefined surveillance goals. First, a disease system model 𝔇 is defined, using observed epidemiologic data and/or theory, and taking into account relevant factors influencing disease dynamics or distribution. Multiple realizations of disease data (*d*) may be generated to explore optimal designs under uncertainty or variability of the underlying system (Section 3.1). Furthermore, an ensemble of disease models can be combined to reduce the chance of model misspecification. Next, a surveillance model is defined to represent how information on the state of the disease system is captured as a function of design parameters *θ* and any other relevant variables (e.g., factors known to affect the sensitivity and specificity of a diagnostic test, or estimated underreporting rates for an area; Section 3.2). To initiate the optimization process, an initial design parameter set, *θ*_1_, is drawn from the design space subject to operational constraints *g(θ*_*i*_*) ≤ 0, h(θ*_*i*_*) = 0* and, along with underlying disease data *d*, input to the surveillance model to generate a realization of surveillance information, 𝒥_1_*=* 𝒥*(θ*_1_, *d)*. The objective function, *f*, is evaluated based on the disease data *d*, and surveillance information 𝒥_1_ (Section 3.3). If a stopping criterion (e.g., reaching a large number of iterations; *de minimis* improvement in objective function) is not met, a new design parameter set, *θ*_*i*_, is proposed from the design space using metaheuristic search algorithms (e.g., simulated annealing, genetic algorithm, particle swarm algorithm) when the design space is large, or enumeration when the design space is small. This new design parameter set is then used to generate a new realization of surveillance information and evaluation of the objective functions (Section 3.4). After a stopping criterion is met, design parameter sets with the best objective function values are output as optimal surveillance designs.

### Box 1. Surveillance System Optimization Procedure

**Input:** Epidemiologic data and/or theory, surveillance performance data and/or theory, and other auxiliary data (e.g., disease risk factors)

**Output:** the design parameter set with the highest/lowest (i.e., optimal) objective function value

**Initialization**

Define a disease system model to represent the underlying dynamics of the target disease system in the spatial, temporal, and demographic context of interest

Generate disease distributions *d* as realization(s) of the system

Sample initial design parameter set, *θ*_1_, within the design space subject to constraints *g(θ*_*i*_*) ≤ 0, h(θ*_*i*_) = 0

Generate realization(s) of surveillance information, 𝒥_1_, given *d* and *θ*_1_

Evaluate objective function(s) *f* given 𝒥_1_ and *d*

**while** stop criterion is not met **do**

Propose a new design parameter set, *θ*_*i*_, within the design space using metaheuristic search algorithms or enumeration

Generate realization(s) of surveillance information, 𝒥_*i*_, given *d* and *θ*_*i*_

Evaluate objective function(s), *f*, given 𝒥_*i*_ and *d*

**end while**

**return** the best design parameter set, 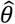 (i.e., with the optimal objective function value)

### 3.1 Specify and parameterize disease system model

An accurate representation of epidemiologic characteristics of the target disease(s) is essential for a successful optimization. This representation can be generated using observational data, outputs of mechanistic transmission models, or other approaches, and represents the best estimate of the disease’s epidemiology that is used to evaluate surveillance network performance using objective functions (defined in section *3.3 Define objective function(s)*, below). To avoid potential model-misspecification, an ensemble of disease models and multiple realizations of disease models (i.e., with varying epidemiologic parameter values) can be utilized in the framework. The structure of the disease system model output—such as spatial and temporal resolution—should be tailored to the surveillance objectives and design parameters. For instance, if a surveillance objective is to better estimate the spatial distribution of a disease, the target disease data must include geographical information about cases.

### 3.2 Specify and parameterize surveillance model

In order to identify optimal network designs, a model representing key aspects of the sampling of and extraction of information from underlying disease processes by the surveillance system is needed. The surveillance model must represent the mechanisms through which variation in network design parameters is expected to impact the epidemiologic information obtained and thus governs optimization with respect to system objectives. Surveillance models generally comprise a set of probability distributions relating target estimands to the underlying disease distribution, conditional on network design and other relevant considerations. For example, to optimize the diagnostic protocol for minimal bias in reporting, a surveillance model may be constructed for the distribution of reported cases conditioned on diagnostic method, background prevalence of the target disease and conditions with similar clinical presentation, and the distribution across subpopulations of factors that impact diagnostic sensitivity and specificity. When random errors contributed by surveillance processes are not explicitly taken into account, as may be the case when seeking to maximize the size of the population covered by a surveillance network, the surveillance model becomes a set of conditional Dirac delta distributions, and is deterministic. During the process of surveillance model specification, aspects of surveillance design that will be allowed to vary during optimization (i.e., the parameters to be optimized), and those that will be fixed (i.e., design aspects that are relevant to performance, but which it is not feasible or desirable to change) must be decided upon. Surveillance models may be as granular (e.g., modeling the full sequence of events necessary for each individual case to be reported) or abstract (e.g., modeling the overall proportion of cases detected in a population) as is deemed necessary for the optimization procedure, recognizing, however, that computational complexity may limit the feasibility of certain representations.

### 3.3 Define objective function(s)

Changes to design parameters can be analyzed in relation to their influence on network performance in the context of specific surveillance system objectives. That is, performance is evaluated with respect to achieving a specific goal or goals. This evaluation is formalized by defining objective functions, which define the specific minimization or maximization problem to be solved, based on the design parameters and surveillance goals of interest. Thus, network performance is estimated through the iterative evaluation of objective functions, which are minimized (or maximized) as the design parameter space is searched. Table 2 presents canonical objective functions available for use in surveillance network optimization. Our examples do not explicitly include operational considerations within objective functions, but these can easily be taken into account. For example, the objective function could be established so as to yield the marginal information gain per added site or sample, or per dollar spent on surveillance.

**Table 2.**
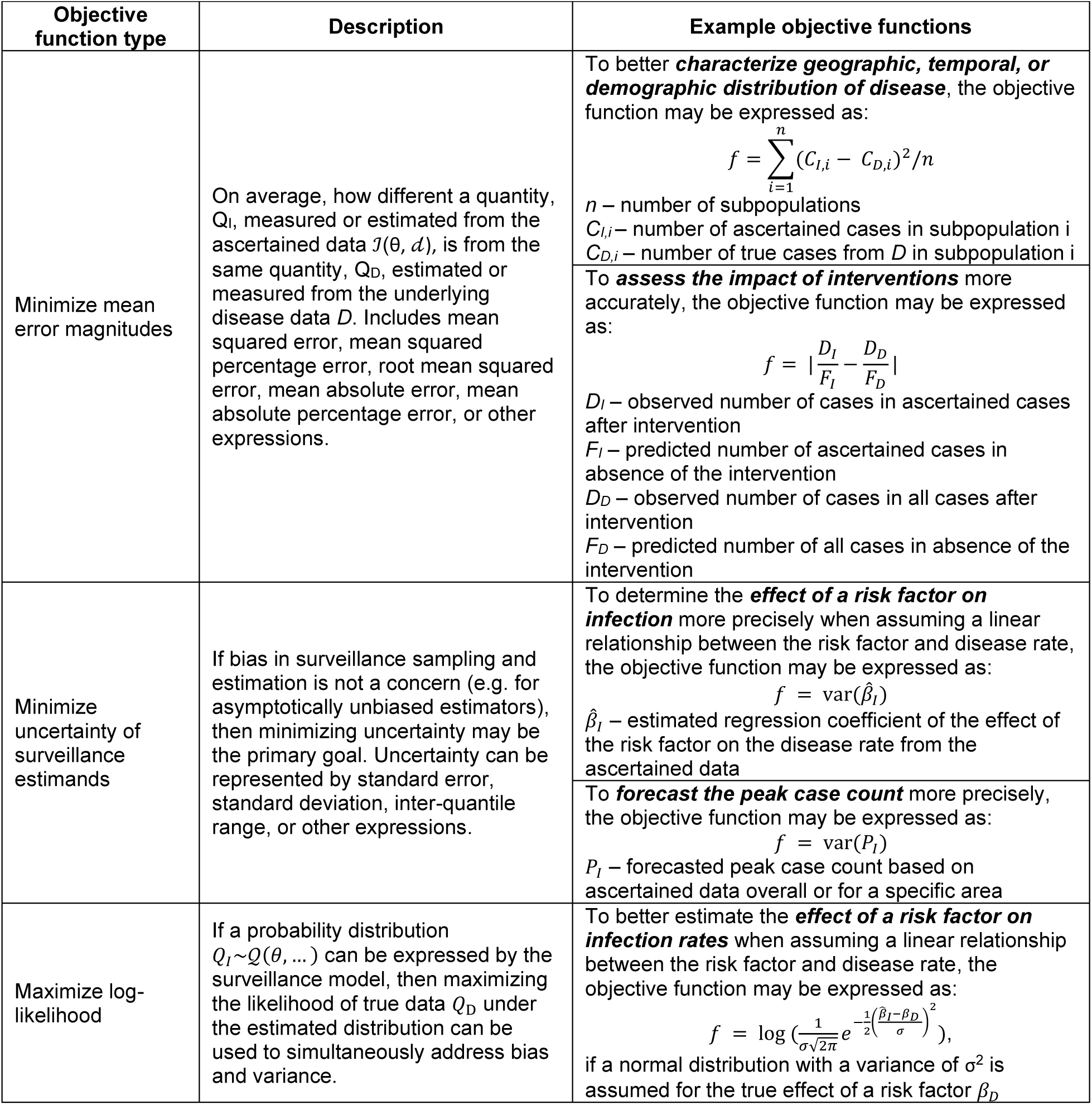
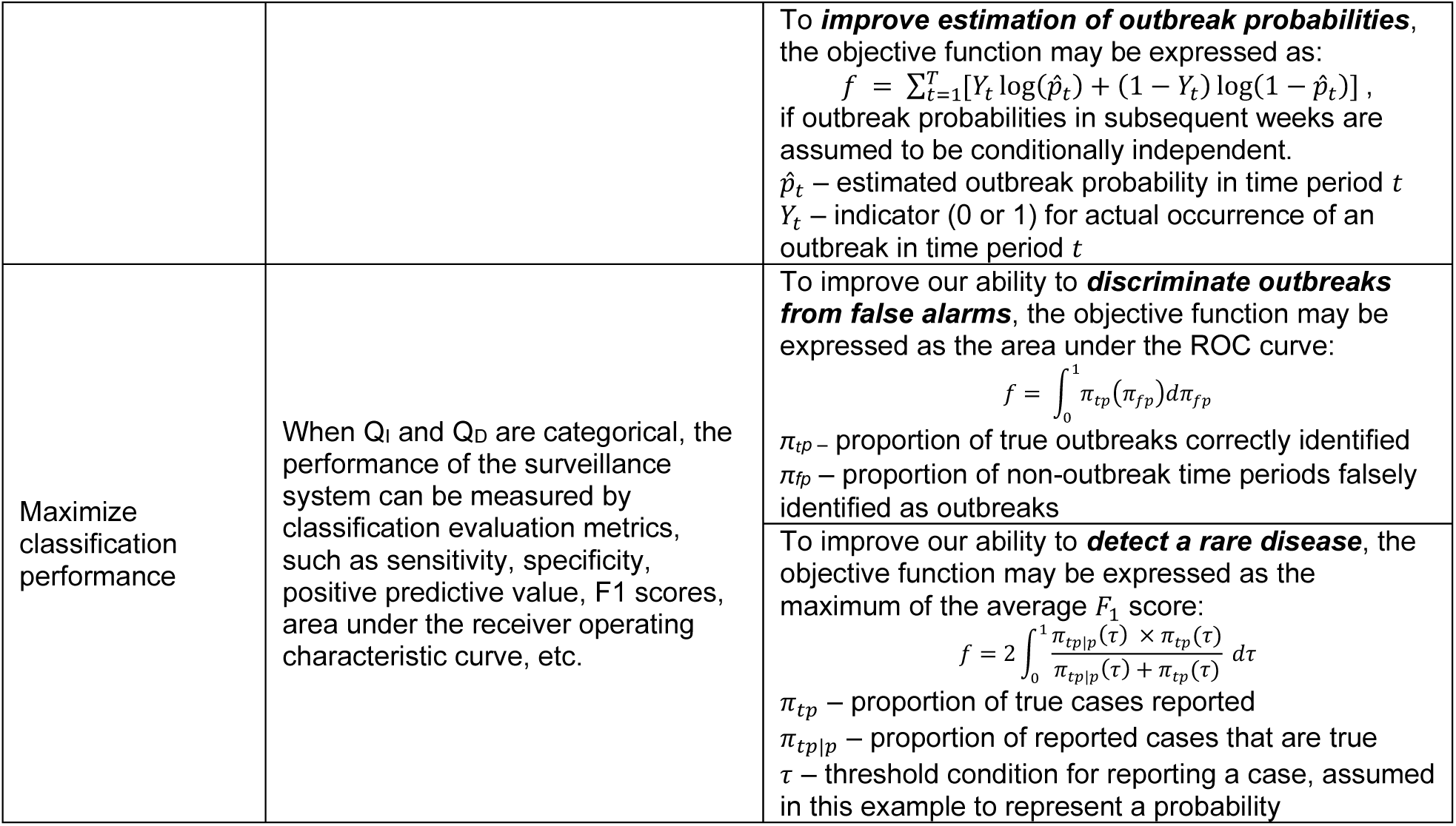
Examples of objective functions for optimization analysis of surveillance networks

### 3.4 Simulation optimization search

The goal of the optimization process (*while* block in Box 1; the loop in *Simulation optimization search* component of Figure 1) is to thoroughly explore the response surface of the objective function(s) over the design space so as to identify designs likely to yield optimal or near-optimal surveillance performance. Candidate surveillance designs are drawn from the design space, and the expectations of resulting objective function values across realizations are evaluated with respect to the simulated true and ascertained disease data; this process is repeated iteratively until a stopping criterion is reached, e.g., the convergence on estimated optimum(a); exhaustive sampling of the design space; or the exceedance of a computational budget. When the design parameter space is small, exhaustive evaluation of objective function values across the entire design parameter space is possible. Sufficient and efficient searching of large design parameter spaces, by contrast, requires heuristic or metaheuristic optimization algorithms (e.g., simulated annealing, genetic algorithms, particle swarm optimization, or Bayesian model based optimization).

Multiple surveillance objectives can be optimized simultaneously through multi-objective optimization approaches, such as through weighted sums of objective functions or Pareto optimization [40]. Generating weighted sums of objective function values allows for the specification of relative importance of different objectives. If one objective is less important, it would be assigned a smaller weight when compared with other objectives. In this way, optimal designs are not overly influenced by less important objectives. Pareto optimization outputs a set of optimal solutions (Pareto optimal set) for which no other solutions can perform better under all objectives. That is, improving the performance on one objective leads to worsening at least one of the other objectives. Decision makers are then tasked with choosing the “best” design from the Pareto optimal set based on other considerations. Multi-objective optimization in the presence of a large design space can be handled by modified metaheuristic algorithms [41]. For example, to accommodate multiple objectives, Pareto simulated annealing approaches seek to express the acceptance probability of a new design as a function of its improvements in all objectives when compared with the current best design [42].

### 4 Demonstration of the surveillance simulation and optimization framework: optimal selection of new surveillance sites

Here, we demonstrate an application of our surveillance system optimization framework in the context of selecting candidate sites to add to an existing cross-sectional survey network. We consider two surveillance design objectives—optimal prediction of the geographical distribution of the disease (hereafter referred to as *spatial prediction*) and optimal estimation of the effect of a risk factor (hereafter referred to as *effect estimation*). We demonstrate how optimal designs can vary in relation to epidemiological characteristics of the target disease; in this case, the rate of decrease in correlation of disease prevalence rates over distance, which determines whether prevalence changes abruptly or smoothly over the spatial domain.

We first describe the demonstration setting, the data available for design optimization, the specification and parameterization of the disease and surveillance system models, and the resulting formalized objective functions for optimizing spatial predictions and effect estimation. We demonstrate the use of an exhaustive search strategy to find the single most optimal site to add to the existing network for both goals, as well as the Pareto-optimal set of single sites to add when considering both objectives simultaneously. We simulate the addition of an arbitrary number of sites, acknowledging that in real-world applications of the framework, the number of sites might be determined by budgetary constraints and/or the marginal informational gains per added site. We conclude our demonstration by considering the best set of three sites to add, which introduces substantial combinatorial complexity, motivating the use of a metaheuristic algorithm to efficiently search for optimal regions of design space.

### 4.1 Demonstration setting

We generated a set of 100 potential surveillance sites scattered uniformly at random across a unit grid, and randomly selected 30 sites to represent a virtual existing surveillance network. We seeded two point sources for a risk factor influencing expected disease prevalence rates (Figure 2A), then simulated disease prevalence under two scenarios of spatial auto-correlation by adjusting the scale parameter (*ρ*) of a log-Gaussian spatial process centered on a linear function of the risk factor. We refer to these as spatially patchy (*ρ = 0*.*1*; Figure 2B) and spatially smooth (*ρ = 0*.*3*; Figure 2C) disease scenarios. Additional details of data generation are provided in Text S1.

**Figure 2.**
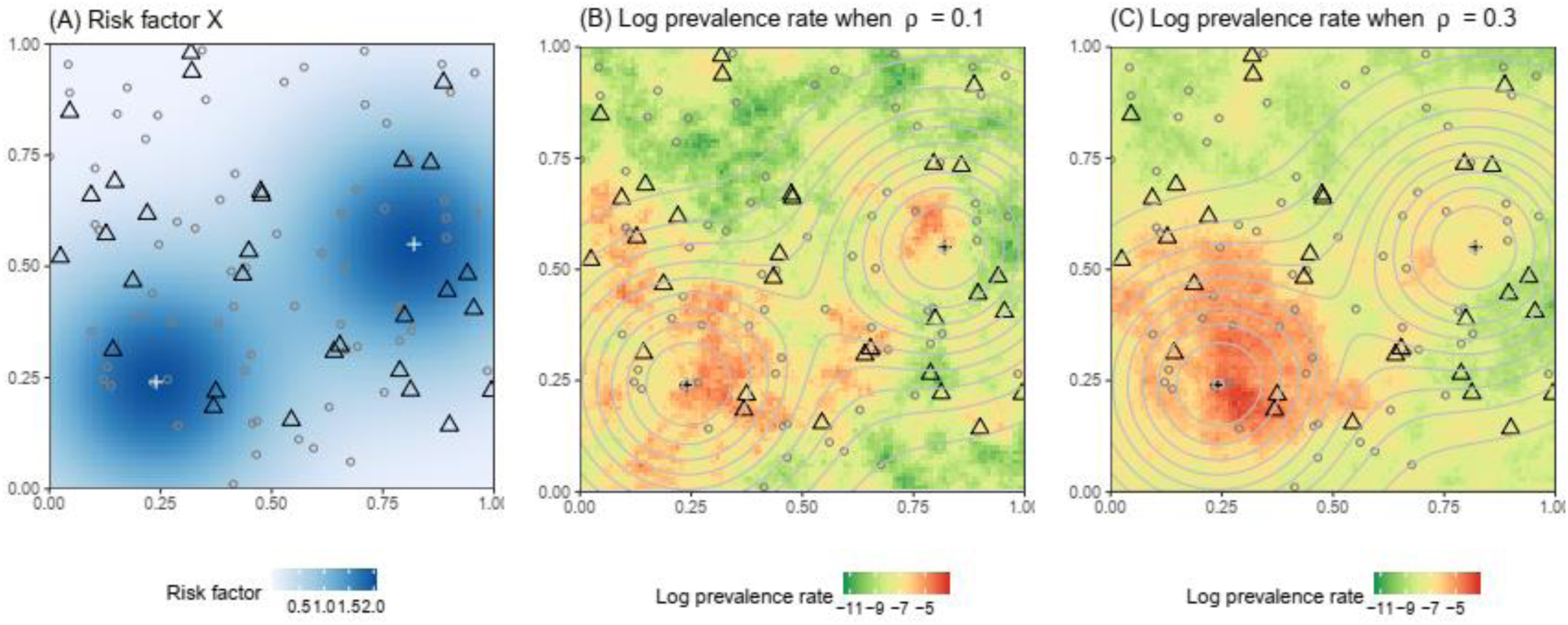
Simulated data used for surveillance system optimization. Spatial variation of (A) the risk factor *X* and (B) log prevalence when ρ = 0.1 and (C) ρ = 0.3. Triangles represent the existing 30 surveyed sites; dots represent the 70 candidate sites; crosses represent two point sources of the risk factor of interest (e.g. locations of mass gatherings); background color in Panel A and contour lines in panels B and C represent the levels of risk factor *X*. Three realizations of the log prevalence surface when ρ = 0.1 or 0.3 are shown in Figure S1.

### 4.2 Data and knowledge inputs

Available epidemiologic data to characterize the relevant aspects of the disease system include simulated prevalence rates observed at the 30 sites enrolled in the surveillance network. Data characterizing the surveillance system and design space include the coordinates of the 30 enrolled and 70 candidate sites. Additional data to support optimization include levels of risk factor *X* at each sampling location. Theoretical inputs include the assumption of a log-linear relationship between *X* and disease prevalence, and that spatial disease prevalence residuals follow a Gaussian process with exponential covariance function.

### 4.3 Set up and initialization

### Disease system model specification and simulation

In this demonstration, relevant aspects of the disease system include the correlation of disease outcomes over space, as well as the association of disease outcomes with risk factor *X*. Based on the observed disease prevalence at participating sites, we assume the log of the prevalence value *Y* is generated from an underlying random spatial process with an *i*.*i*.*d* mean-zero normally distributed noise *ε* with a variance of *σ*_*d*_^*2*^, and can be modelled by:

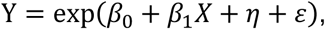

where *β*_0_ represents log of the overall mean prevalence rate, *β*_1_represents the effect of a unit increase in risk factor *X*, and *η* represents a mean-zero Gaussian process accounting for spatial correlation induced by additional dependence not captured by *X*. The spatially correlated error term *η* follows a multivariate normal distribution with a variance-covariance matrix ***C***, in which each entry *c*_*ij*_represents the covariance between the residuals at the *i*th and the *j*th location when *i ≠ j*, and the variance of the residuals at the *i*th location when *i* = *j*. Covariance between sites *i* and *j* is specified 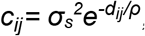, where *d*_*ij*_ is the distance between sites *i* and *j*, and *ρ* is the scale parameter as before; and the variance at site *i* is *σ*_*d*_^*2*^ + *σ*_*s*_^*2*^. The correlation of the residuals between two sites as a function of the distance between them is shown in Figure S1. Parameters *β*_0_, *β*_1_, *σ*_*s*_, *σ*_*d*_, and *ρ* were estimated based on the prevalence rates and risk factor levels at the 30 in-network sites, after which 1000 realizations of log-prevalence rates at the 70 candidate sites were drawn according to the fitted parameters, observed prevalence at in-network sites, risk factor levels at candidate sites, and distance matrix between in-network and candidate sites.

### Surveillance model specification

Relevant aspects of information captured by the surveillance system in this demonstration pertain to the extrapolation of prevalence from enrolled to unenrolled sites, as well as the variance in *X* at enrolled sites. Assuming perfect enumeration of disease prevalence at each enrolled site, as well as known values of the risk factor *X* for all sites, information drawn by each candidate design is represented by improvements in estimates of *β*_1_ and predictions at 70-n out-of-network sites obtained by fitting a universal kriging predictor to data from enrolled sites [43].

### Design space

In our hypothetical example, we have an existing network of 30 surveillance sites {*s*_1_ *… s*_30_}, and 70 additional locations {*s*_31_ *… s*_100_} from which we may select *n* new sites to be added to the network. Therefore, our design parameter *s*_*θ*_is the set of *n* sites to be added to the network, and the discrete design space is all possible sets of *n* sites chosen from 70.

## 4.4 Optimization

### Objective functions: Spatial interpolation

The first surveillance function we wish to optimize is prediction of the geographical distribution of the disease. Therefore, we define the objective function as the mean squared error (MSE) of log predicted prevalence at the *70-n* out-of-network locations after adding *s*_*θ*_ to the network:

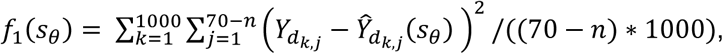

where 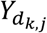 represents the log prevalence rate at the *j*th unenrolled site in the *k*th disease system model realization, while 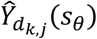 represents the predicted log prevalence rate at the *j*th site after augmenting the existing network with *s*_*θ*_ in the *k*th realization. We denote the objective function value for this objective as OFV1. Other objective functions, such as the MSE of log predicted prevalence rate at the existing 30 sites or across all 100 sites, can also be used. Existing literature on optimal spatial design provides more options for relevant objective functions [44-46].

### Objective functions: Effect estimation

Our second surveillance goal is precise estimation of the effect of the risk factor *X* on the disease outcome, so the objective function is formalized as:

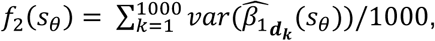

where 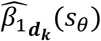 represents the estimate of *β*_1_ after augmenting the existing network with *s*_*θ*_ in the *k*th disease system model realization.

### Search algorithms

When a single site is to be added to the network, the design space is small enough to allow for enumeration of objective function values at all possible designs. Therefore, the algorithm for proposing new designs simply steps sequentially through sites {*s*_31_ *… s*_100_}. However, when the optimization question is shifted to the best three sites to add, the design space expands to 54,740 combinations, making sequential enumeration a prohibitively expensive search strategy. Under these conditions, heuristic (greedy) or metaheuristic algorithms play an important role in finding the optimal or near-optimal solution within a reasonable amount of time [47]. Moreover, the evaluation of objective function values across realizations can be paralleled to further reduce computational time.

We illustrate the use of a simulated annealing (SA) meta-heuristic algorithm popular in spatial sampling network design [48, 49] to more efficiently explore the design space when three sites are to be added. In SA, a random initial design is proposed, after which, at each iteration, a new design is sampled from the neighborhood of the current design and the objective function value (OFV) for the new design is evaluated. Here, the neighborhood of a set of *n* sites to enroll is defined as designs sharing *n-1* sites with the current design. If the new OFV is superior to the current OFV, the new design is accepted as the next design; otherwise, it is accepted with a probability of 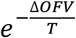, where *ΔOFV* is the change in the OFV and *T* is a tuning parameter analogous to temperature [50]. *T* decreases at a rate *α* after each iteration, causing SA to accept deterioration in the OFV more frequently at the beginning of the run and rarely towards the end. Probabilistically accepting worse solutions early in the search enables the algorithm to escape local optima. For our demonstration, we set the initial temperature *T*_0_ and cooling rate *α* separately for each objective and epidemiologic scenario, following suggestions by Sait and Youssef [50], and set the stopping criteria is to be *T≤10*^*-6*^. We repeat the SA process 3 times to examine the convergence of the result.

## 4.5 Optimal surveillance designs

### Selecting one additional site to optimize spatial prediction

The mean squared error of spatial predictions across unenrolled sites (OFV1) is minimized by enrolling sites that are in close proximity to multiple out-of-network sites — especially clusters of unmeasured sites at long distances from existing network locations (Figure 3, panels A and B). These optimal placements address informational gaps by enrolling sites that increase the average covariance between measured and unmeasured locations, and tend to fall in areas close to several unenrolled sites but away from the initially enrolled locations. Furthermore, the amount of information that can be inferred from the same set of neighboring sites increases with the scale parameter *ρ*. Thus, in the spatially patchy disease scenario, where the scale of spatial autocorrelation is small, optimal placement occurs in the center of a tight cluster of unenrolled sites (Figure 3, panel A). Under the spatially smooth scenario, the same cluster is correlated with initially enrolled sites, and optimal site placement falls in the center of a loose cluster of unmeasured sites located quite far from the initial network (Figure 3, panel B). Under the parameter values used to generate demonstration data, there is no clear influence of risk factor level *X* on site selection to optimize spatial prediction.

**Figure 3.**
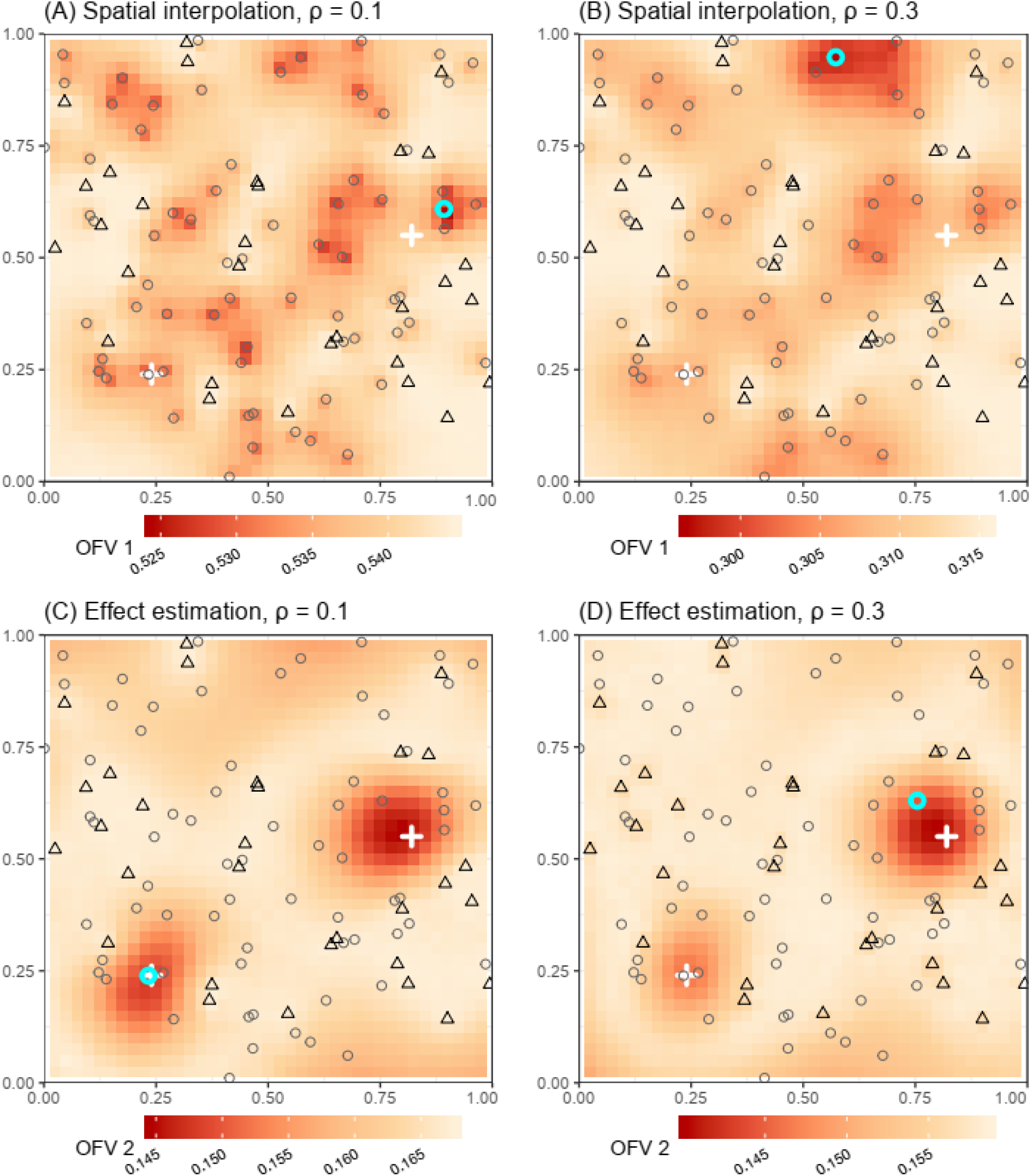
Optimal site placement to augment a surveillance network for spatial prediction or effect estimation under scenarios of spatially patchy or smooth disease distributions. Black triangles represent initially enrolled sites, gray circles represent unselected candidate sites, and the cyan circle indicates the optimal site to add to the network. White crosses represent point sources for risk factor *X*. Raster colors represent objective function values for hypothetical sites added across a regular 41*41 grid in order to visualize the response surface in relation to initial network locations and the underlying risk factor.

### Selecting one additional site to optimize effect estimation

The variance of the effect of risk factor *X* on log disease prevalence (OFV2) is lower when the value of *X* at the cell to be added lies towards an extreme of *X*’s observed range and when the site to be added is relatively uncorrelated with (i.e., distant from) initially enrolled sites (Figure 3, panels C and D). In the spatially patchy disease scenario, where the scale of spatial autocorrelation is limited, optimal site placement is dominated by the level of risk factor *X*, and the available site with highest *X* is chosen (Figure 3C). In the spatially smooth scenario, with an extended scale of spatial autocorrelation, the correlation of outcomes between the site with the highest *X* level and nearby initially enrolled sites results in selection of an alternative location where the value of *X* is less extreme, but prevalence is expected to be more independent of previously observed outcomes (Figure 3D).

### Single site selection based on multiple objectives

When simultaneously optimizing site enrollment for spatial prediction and effect estimation, the output is a Pareto optimal set containing designs that are considered equally optimal because no objective function value can be improved without impairing the other objective function values. A set of six candidate sites emerges for the spatially smooth disease scenario, including four alternative selections to the optimal locations for each single objective (Figure 4). The Pareto optimal set for the spatially patchy scenario includes only one non-dominated site in addition to the optimal locations for either objective individually (Figure S2). Since the solution given by Pareto optimization is not unique, some way of reconciling the objective criteria, such as a weighted sum or expression of total cost may be required to choose the optimal design. Notably, we did not incorporate cost associated with adding sites in our analysis, but this could be accomplished by including a third objective function representing the marginal information gain per added site. In this case, the spatial prediction OFV, effect estimation OFV, and the cost-effectiveness OFV would be jointly optimized.

**Figure 4.**
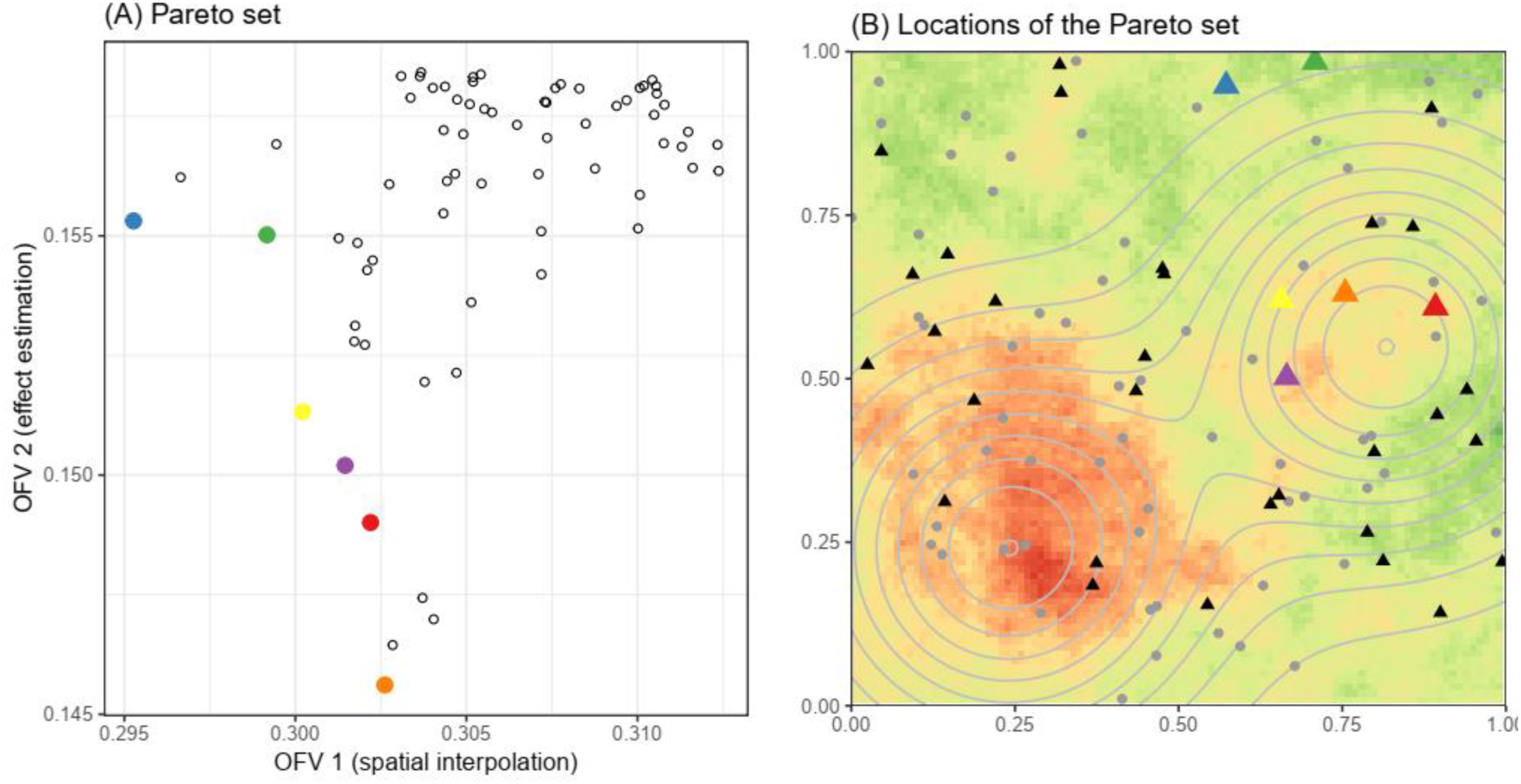
Results from Pareto optimization under the spatially smooth disease scenario (ρ=0.3). (A) Mean squared error of log predicted disease prevalence (OFV1) and variance of causal effect estimate (OFV2) of the Pareto set (colored dots) and all other candidate sites (hollow dots). (B) Locations of the Pareto set (colored triangles) colored coded as in Panel A. Black triangles represent initially enrolled sites, and gray dots represent unchosen candidate sites. Background color in Panel B represents log prevalence when ρ = 0.3 using the same color scheme as in Figure 2C, while contour lines represent levels of risk factor *X*.

### Selecting three additional sites to optimize spatial prediction

As a final example, we demonstrate the use of metaheuristic algorithms to search larger design spaces, applying simulated annealing to select three additional sites out of seventy candidate sites simultaneously. Simulated annealing optimizations seeded with different initial designs converged to the same best set of three additional sites to enroll for enhanced spatial prediction under the spatially smooth disease scenario (Figure 5). All three SA runs (Figure 5A, colored lines) converged to the same optimal design within 6,000 iterations. Given the parameters and the stopping criteria we used, each run terminated after 8,630 iterations. Even with three runs, the total number of objective function evaluations was 25,890, less than half of what would be required if using enumeration. Figure 5B shows the location of the optimal three-site set. The results effect estimation, as well as for the spatially patchy outcome scenario are shown in Figures S3-5.

**Figure 5.**
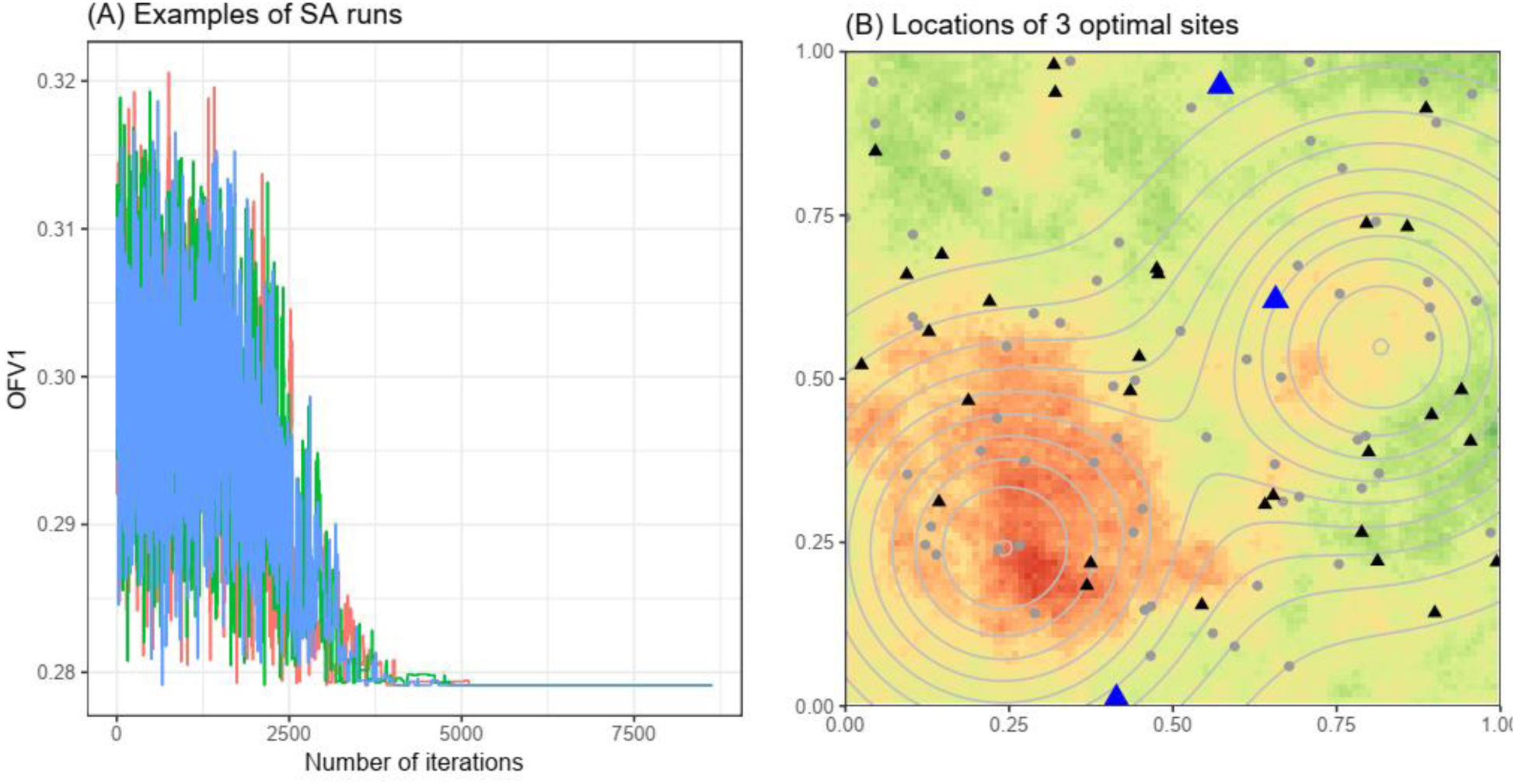
Metaheuristic optimization with simulated annealing (spatial prediction, spatially smooth disease scenario). (A) Mean squared error of predicted log prevalence (OFV1) across iterations of three SA runs. (B) The locations of the optimal 3 sites. Black triangles represent existing sites, blue triangles represent the optimal additional sites, and gray dots represent unchosen alternative sites. Background color in Panel B represents log prevalence when ρ = 0.3 using the same color scheme as in Figure 2C, while contour lines represent levels of risk factor X.

## 5. Conclusion

Surveillance system designs that provide reliable, timely estimates of the spatial-temporal distributions of endemic and epidemic diseases, are critical to the efficient allocation of resources for public health responses. However, opportunities to apply numerical optimization to surveillance system design have heretofore been overlooked in the literature. In this paper, we have presented a framework for surveillance optimization via simulation to enhance design decision making and facilitate research into optimal design principles under uncertain or changing epidemiological conditions. While we focus on surveillance of human disease, the framework could also be applied to the optimization of vector or environmental surveillance.

The framework presented can arrive at improved surveillance system designs through the incorporation of data and models of local disease transmission status, diverse surveillance goals, resource and operational constraints, and by stimulating collaboration between health planners, researchers, and software developers. However, it should also be recognized that the rationality of the output optimal design will be highly dependent on the accuracy and relevance of data or models used to represent disease and surveillance processes during optimization, as well as the performance of the optimization search algorithm. There is much future work to be done to develop and validate simulation models that can represent relevant case generating and measurement processes accurately; to analyze the sensitivity of optimal design to the specification of disease system models and changes in disease epidemiology; and to adopt optimization approaches from related fields—such as environmental monitoring network design and signal processing [51-53]—to disease surveillance design applications.

## Data Availability

Not applicable

